# Are specific, measurable, action-oriented, realistic, and time-bound (SMART) goals associated with improved walking outcomes for stroke survivors undergoing outpatient stroke rehabilitation? An observational cross-sectional retrospective cohort study

**DOI:** 10.1101/2024.09.05.24313054

**Authors:** Helia Mohammadi-Ghayeghchi, Nidhi Bhagat, Mackenzie Campbell, Madi Mayhew, Rheall Dufresne, Levi Ewald, Augustine J. Devasahayam, Avril Mansfield

## Abstract

**Background:** Goal-setting is a core principle used in clinical practice to guide treatment. Setting goals improves adherence to rehabilitation treatments and may lead to better outcomes in people with neurological disorders. However, there is a lack of research into the prevalence of goals using a Specific, Measurable, Action-Oriented, Realistic, and Time-Bound (SMART) framework. Additionally, it is currently unclear if the SMART framework improves ambulatory outcomes in outpatient stroke rehabilitation.

**Methods:** This observational, cross-sectional, retrospective cohort study reviewed charts of all patients admitted to outpatient stroke rehabilitation at three hospitals over a 1-year period. Patients were included in the analysis if they had documented ambulatory goals. Goals were classified as either SMART or non-SMART. Analysis of covariance was used to compare Functional Ambulation Category scores at discharge between the SMART and non-SMART groups, controlling for admission scores, length of stay, and time post-stroke.

**Results:** 300 patients were included in the analysis. Of these, 61 (20.3%) had at least one SMART ambulatory goal. Analysis of covariance revealed a statistically significantly greater Functional Ambulation Category scores at discharge for the SMART goal group compared to the non-SMART group (mean Functional Ambulation Category scores at discharge [95% confidence interval], SMART group: 4.2 [4.0, 4.5], non-SMART group: 3.8 [3.6, 4.1]; F_1,60_ = 4.40, p = 0.043).

**Conclusion:** The use of SMART goals in outpatient stroke rehabilitation is associated with better ambulatory outcomes compared to non-SMART goals. These findings suggest that incorporating the SMART framework in clinical practice can enhance the effectiveness of rehabilitation interventions for stroke patients. Further studies are recommended to explore the long-term effects and broader applications of SMART goal-setting.

## INTRODUCTION

Stroke is a major cause of disability globally (1, 2). The economic burden from stroke and its resulting long term disability is significant; in Canada, the overall direct healthcare costs in the first year after an individual experiences stroke are approximately $60,658 (2, 3). However, with the implementation of organized stroke care delivery and evidence-based guidelines, such as *The Canadian Stroke Best Practice Recommendations*, improvements in outcomes and reduced economic burden have been reported (4-6). Currently, 80% of individuals survive strokes and live with its effects, and the total number of Canadians living with disabilities resulting from stroke is expected to double within the next 20 years (7).

While the effects of stroke vary depending on the severity and location of the stroke, gait impairments are common and experienced by approximately 78% of stroke survivors (8).

Improving ambulatory ability is one of the top priorities for stroke survivors as it impacts their quality of life and long-term health (9, 10). As such, a large component of stroke rehabilitation involves improving gait, and ambulatory goals are often the most commonly reported rehabilitation goals post-stroke (9, 11-13). According to the current *Canadian Stroke Best Practice Recommendations* (4), rehabilitation should be “goal-oriented”. Goal-oriented care can better motivate individuals, guide the therapeutic intervention, and promote patient-centered care as patients work towards improving ambulation (4, 10). The SMART framework, which grew from business culture, provides a structure for setting health-related goals (14).

It is unclear how frequently clinicians apply standardized approaches, such as the SMART framework, to goal-setting in rehabilitation practice. The existence of multiple variations of the SMART acronym in the literature (15), and the time-intensive nature of creating SMART goals can negatively impact their usage. In a previous study, only 41% of documented goals within inpatient stroke rehabilitation adhered to the SMART criteria, with the timeline being the least frequently documented component (16). While standardized goal-setting like the SMART framework is thought to contribute to increased rehabilitation participation and task performance (17, 18), its benefits may be limited if SMART goals are not widely adopted within practice. Since nearly 60% of stroke survivors are referred to outpatient stroke rehabilitation after inpatient rehabilitation (19), it is important to determine the prevalence of goals that follow the SMART framework in outpatient settings.

While the SMART framework offers a structured approach to goal-setting (10, 20), it is also unclear if SMART goal-setting in rehabilitation improves patient outcomes (21). Specifically, no studies to date have investigated the effects of SMART goal-setting on walking outcomes in the stroke population. Improving ambulatory ability through stroke rehabilitation would not only positively impact the stroke survivor’s quality of life (9, 10), but it would also reduce the economic burden associated with stroke, as a large proportion of the cost of stroke is directly due to the stroke survivors’ inability to walk independently (22). Therefore, understanding whether SMART goals result in improved ambulatory outcomes in stroke survivors attending stroke rehabilitation is crucial to determine the efficacy of this framework in this setting.

The primary objective of this study was to determine the prevalence of ambulation-related SMART goals that adhere to the SMART framework among stroke patients in the outpatient setting. The secondary objective was to determine whether patients who set SMART ambulation goals at admission have improved Functional Ambulation Category (FAC) scores at discharge compared to those with non-SMART goals. We expected that 41% of ambulatory goals in outpatient rehabilitation would adhere to SMART criteria, based on prior research conducted within inpatient rehabilitation (16). Additionally, the mean change in FAC scores following outpatient rehabilitation was expected to be significantly greater in stroke survivors with SMART ambulatory goals compared to those without SMART ambulatory goals.

## METHODS

### Study design

This observational cross-sectional retrospective cohort study involved secondary analysis of data collected as part of a mixed-methods study (23, 24) at three urban rehabilitation hospitals in Toronto: Toronto Rehabilitation Institute – University Centre site, Toronto Rehabilitation Institute – Rumsey Centre site, and St. John’s Rehabilitation – Sunnybrook Health Sciences Centre. All hospitals have dedicated outpatient stroke rehabilitation programs. The study was approved by the University Health Network (protocol number: 20-5695) and Sunnybrook Health Sciences Centre (protocol number: 36050) Research Ethics Boards, with a waiver of patient consent for inclusion in the chart review approved.

### Participants and data collection

Patients were included in the chart review if they: were admitted to one of the three sites during a 1-year period following 2021; had a confirmed stroke; and had at least one ambulation-related goal documented in their chart.

The FAC was used to evaluate patients’ ambulatory capability by assessing the level of physical support required when walking short distances and over various surfaces (25, 26). This measure is commonly used in stroke care and research, as it has good responsiveness to gait-related changes within the first few weeks and after 6 months of rehabilitation (26). In addition, it has high test-retest reliability (ϰ = 0.950), interrater reliability (ϰ = 0.905), and concurrent validity with significant correlation with results of the 6-minute walk test, walking velocity, and stride length (26). FAC scores for admission and discharge were determined based on clinical notes written by the clinician, and were available for the majority of patients. FAC scores and SMART ambulatory goals (see below) were determined by two different sets of researchers independently. All patient goals within the chart abstraction forms were extracted verbatim as documented in the patient charts, omitting any potentially identifying information.

Patient age, sex, location and type of stroke, and affected side of the body were extracted from patient charts as cohort descriptors.

### SMART ambulatory goal classification

Ambulation-related goals were defined as goals that related to improving gait with or without a gait aid/human support; included the words “walk”, ambulate”, or “gait” relating to the patients’ ability to mobilize; related to activities or tasks where walking is a major component (e.g., hiking, going up or down stairs); or were related to elements of the gait cycle (e.g., circumduction, hip hike). Patient charts were divided between two rating groups, each consisting of 2 authors, for identifying whether the patient had ambulatory goals for inclusion in the study. Within each group, charts were reviewed independently by both researchers and rated as ambulatory or non-ambulatory. Inter-rater agreement of the definition of ambulation-related goals was conducted using Fleiss’ kappa. Fleiss’ kappa of 0.978 was achieved, indicating almost perfect agreement (25). Patients were determined to have an ambulatory goal, and subsequently included in the study, if both raters agreed that the patient had one or more goals in their chart that met one of the above ambulatory criteria. In cases of disagreement between the two raters, the goals were rated independently by a third group of two authors. If disagreement persisted between raters, the goals were discussed among all 6 raters until consensus was achieved.

Patients were considered to have SMART ambulatory goals if they had at least one ambulatory goal that met all 5 SMART criteria. For the purpose of this study, the SMART components were specific, measurable, action-oriented, realistic, and time-bound (27). Goals were action-oriented if they centred around executing a particular action; since all goals were focused on the action of walking, all goals were considered action-oriented. We were unable to evaluate if goals were realistic based on information presented in the charts, and therefore assumed that physiotherapists only documented goals that they believed to be realistic. Therefore, we judged if goals met three of the SMART criteria: specific, measurable, and time bound. Goals were considered to be specific if the intended achievement or result, such as endurance, coordination, gait quality/pattern, speed, distance, etc., in relation to ambulation was clearly specified and the conditions of performance were described, including elements such as the amount needed to improve, level of assistance, the use or absence of gait aids, and the environment. Goals were considered measurable if they enabled objective assessment of progress and/or the goal and progress could be monitored through outcome measures, or explicit/implicit observation. Goals were time-bound if they were structured with a deadline, indicated by a specific time point (e.g., an event like discharge or a particular date). The definitions of all 5 letters within the acronym were based on the definition provided by the College of Physiotherapists of Ontario, and adapted by the authors to be specific to ambulatory goals (27).

Prior to categorizing goals, inter-rater agreement of the SMART criteria was determined using Fleiss’ kappa to ensure uniformity in the application of the SMART criteria to ambulatory goals. Fleiss’ kappa was 0.868, which indicates an almost perfect interrater agreement (28). As above, goals were classified as SMART or non-SMART by two groups of raters independently.

### Statistical methods

Descriptive statistics were performed for the cohort descriptors. The Shapiro-Wilk test was used to test normality of continuous variables (age, length of stay, time post stroke). To address the primary objective, the prevalence of patients with SMART ambulatory goals within the cohort was obtained by dividing the total number of individuals with at least one SMART ambulatory goal by the total number of individuals within the cohort. To address Objective 2 and determine whether patients who set SMART ambulatory goals at admission have improved FAC scores at discharge compared to patients with non-SMART ambulatory goals, an analysis of covariance (ANCOVA) was used to compare FAC scores at discharge between SMART and non-SMART groups while controlling for FAC scores on admission, length of stay and time post-stroke. Length of stay was controlled for as longer lengths of stay may indicate greater therapy time, and a positive dose-response relationship between time scheduled in therapy and improvements in clinical outcomes has been demonstrated (29). Time post stroke is expected to impact the extent to which the patient can improve as there is a critical period post stroke in which patients are more responsive to rehabilitation (30). All statistical analyses were conducted using Stata (Stata Statistical Software: Release 18, StataCorp LLC, College Station, Texas, USA). Stacked proportional bar graphs (‘Grotta bars’), controlling for confounding variables (FAC admission scores, time post-stroke, and length of stay) were created to visualize the data (31, 32) using R (version 4.4.1, R Foundation for Statistical Computing, Vienna, Austria).

### Sample size justification

604 patients were admitted to the three hospitals during the review period. A previous study found that 62% of patients in outpatient stroke rehabilitation have ambulatory goals (12). Therefore, we expected to include 62% of patients, or 375 patients, in the analysis. A sample size of 375 would allow us to estimate, with 95% confidence, an expected SMART goal prevalence of 41% with a precision of ± 5% for Objective 1. For Objective 2, a sample size of 375 would provide >99% power to detect a difference of 0.5 in FAC between the SMART and non-SMART, assuming 41% of patients are in the SMART group and three covariates (33).

## RESULTS

Of 604 patients who were admitted to the outpatient units, 7 were excluded because they were missing key information (e.g., stroke date), 58 were excluded because they had no documented rehabilitation goals, and 239 were excluded because they did not have any ambulatory rehabilitation goals. This left 300 patients for inclusion in the study. Patient characteristics are reported in Table 1.

**Table 1:** Participant characteristics. Values presented are means with standard deviations in parentheses for continuous variables, or counts with percentages in parentheses for count variables. Percentages might not sum to 100% due to rounding error.

|  | Facility 1<br>(n=126) | Facility 2<br>(n=63) | Facility 3<br>(n=111) |
| --- | --- | --- | --- |
| Age (years) | 68.6 (12.1) | 63.4 (13.3) | 65.0 (14.4) |
| Sex, n women (%) | 44 (34.9) | 29 (46.0) | 49 (44.1) |
| Time post-stroke (days) | 96.6 (90.6) | 142.9 (176.1) | 91.1 (57.3) |
| Stroke type, n (%) |  |  |  |
| Ischemic | 95 (75.4) | 45 (71.4) | 92 (82.9) |
| Lacune | 2 (1.6) | 2 (3.2) | 3 (2.7) |
| Hemorrhage/hemorrhagic | 20 (15.9) | 13 (20.6) | 10 (9.0) |
| Not available | 3 (2.4) | 0 (0) | 0 (0) |
| Other | 6 (1.8) | 3 (4.8) | 6 (5.4) |
| Affected hemisphere, n (%) |  |  |  |
| Left | 52 (41.3) | 36 (57.1) | 48 (43.2) |
| Right | 59 (46.8) | 23 (36.5) | 51 (46.0) |
| Both | 15 (11.9) | 4 (6.4) | 11 (9.9) |
| Not available/unknown | 0 (0) | 0 (0) | 1 (0.9) |
| Affected side of body, n (%) |  |  |  |
| Left | 54 (42.9) | 20 (31.8) | 52 (46.9) |
| Right | 55 (43.7) | 36 (57.1) | 47 (42.3) |
| Both | 14 (11.1) | 3 (4.8) | 10 (9.0) |
| Neither | 3 (2.4) | 4 (6.4) | 2 (1.8) |
| Length of stay (days) | 73.8 (35.6) | 104.7 (52.3) | 93.6 (51.3) |
| SMART goals, n (%) | 65 (51.6) | 0 (0) | 0 (0) |
| Specific goals, n (%) | 123 (97.6) | 50 (79.4) | 84 (75.7) |
| Measurable goals, n (%) | 105 (83.3) | 47 (74.6) | 72 (64.9) |
| Time-bound goals, n (%) | 77 (61.1) | 0 (0) | 1 (0.9) |

Regarding Objective 1, 61/300 patients (20.3%; 95% confidence interval: [15.7, 24.9]%) had at least one SMART ambulatory goal; this proportion was significantly below the expected proportion of 41% of patients with SMART ambulatory goals.

There were no patients in Facility 2 or 3 with SMART ambulatory goals. Therefore, analysis for Objective 2 only included patients from Facility 1. Of the 126 patients at Facility 1, 12 were excluded from analysis of Objective 2 because they were missing FAC scores at admission and/or discharge, and 49 were excluded because they had FAC scores of 5 on admission, leaving 65 patients for analysis. FAC scores on admission and discharge from outpatient rehabilitation for these 65 participants are presented in Table 2. When controlling for length of stay, time post-stroke, and FAC scores on admission, there was a statistically significant difference in FAC scores at discharge between the SMART and non-SMART groups (mean FAC scores at discharge [95% confidence interval], SMART group: 4.2 [4.0, 4.5], non-SMART group: 3.8 [3.6, 4.1]; F_1,60_ = 4.40, p = 0.043). There were more patients with discharge FAC scores of 3 and 5 in the SMART group than the non-SMART group (Figure 1). Additionally, there were 3 patients in the non-SMART group with discharge FAC scores of 0 or 1, compared to no patients in the SMART group.

**Table 2:** FAC scores on admission and discharge. Values presented are the number of patients within in each functional ambulation category at admission and discharge for patients included in analysis of Objective 2. Discharge numbers are unadjusted for covariates; adjusted values are presented in Figure 1.

| FAC | SMART group<br>(n=34) |  | Non-SMART group<br>(n=31) |  |
| --- | --- | --- | --- | --- |
|  | Admission | Discharge | Admission | Discharge |
| 0 (Non-ambulatory) | 2 | 0 | 2 | 1 |
| 1 (Continuous manual assistance) | 1 | 0 | 3 | 2 |
| 2 (Light touch assistance) | 4 | 1 | 4 | 3 |
| 3 (Standby guarding) | 8 | 7 | 9 | 5 |
| 4 (Supervision in some circumstances) | 19 | 5 | 13 | 9 |
| 5 (Independently ambulatory everywhere) | 0 | 21 | 0 | 11 |

**Figure 1:**
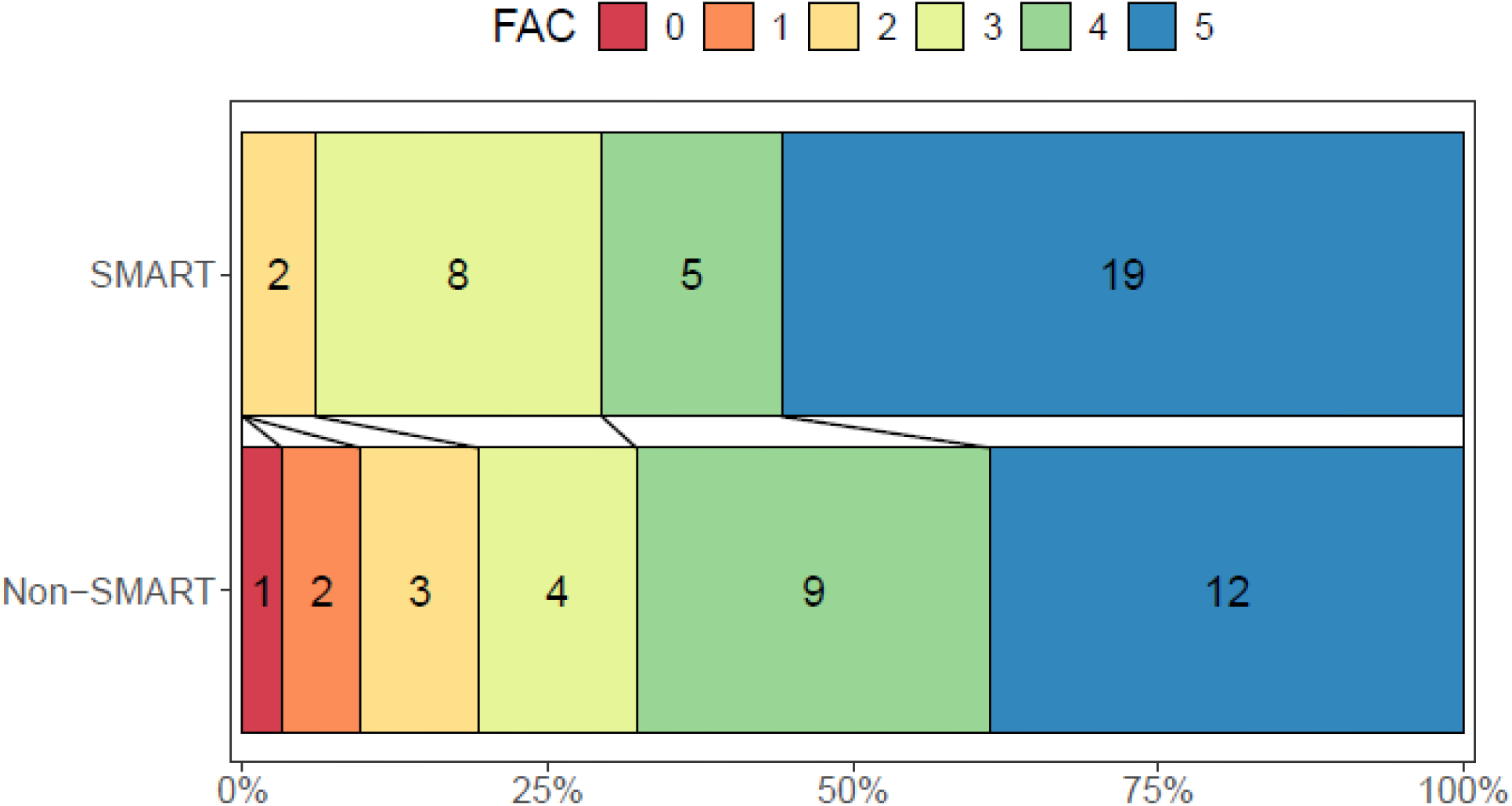
Stacked proportional bar graphs (Grotta plots) comparing Functional Ambulation Category (FAC) scores between groups at discharge. Plots show the number of patients in each category, controlling for confounding variables (FAC scores on admission, length of stay, and time post-stroke (31)).

## DISCUSSION

The objectives of this study were to determine the prevalence and impact of SMART goals on ambulatory outcomes in outpatient stroke rehabilitation. The study revealed that only 20.3% of patients had SMART ambulatory goals, highlighting a limited adoption of the SMART framework during goal-setting in outpatient stroke rehabilitation. Despite the relatively low use of SMART goals, the findings indicate that patients with SMART ambulatory goals had statistically significantly higher FAC scores at discharge from outpatient stroke rehabilitation compared to those with non-SMART goals.

The low prevalence of SMART goals in rehabilitation settings found in this study are in line with those found in a study observing inpatient stroke rehabilitation, in which a 41% prevalence of SMART goals was observed (16). The noticeably lower prevalence determined in the current study compared to the 2018 study by Plant and Tyson may be due to the fact that these studies were conducted within the context of different healthcare systems. The study by Plant and Tyson was conducted in the United Kingdom. Stroke care in the United Kingdom is guided by the recommendations set out by the *National Institute for Health and Care Excellence* (34). In addition to stating that stroke rehabilitation should be goal-oriented, these guidelines provide explicit criteria that goals should meet. According to *National Institute for Health and Care Excellence* (34), clinicians should ensure that stroke survivors have rehabilitation goals that are meaningful and relevant, activity-focused, challenging but achievable, and that they include short term and long term elements. These guidelines follow to a large extent the SMART acronym used in the Plant and Tyson study (specific, measurable, achievable, realistic, timely) (16). Since goal-formatting is clearly described by the *National Institute for Health and Care Excellence*, it is likely that more clinicians practicing in stroke rehabilitation in the United Kingdom would follow these guidelines, ultimately creating goals that would be captured as SMART. In contrast, despite indicating that rehabilitation and tasks should be goal-oriented, the *Canadian Stroke Best Practice Recommendations* do not specify specific criteria that stroke survivor’s goals should meet (4). Thus, clinicians practicing in stroke rehabilitation in Canada may be less likely to format their goal following the SMART framework as it is not indicated in the recommendation. Therefore, the study conducted by Plant and Tyson likely saw a greater prevalence in the adoption of SMART goals due to this difference in goal setting recommendations between the two healthcare systems. The addition of such criteria for goal setting within the *Canadian Stroke Best Practice Recommendations* may increase the use of SMART goals within stroke rehabilitation in Canada.

Another reason for the low prevalence of SMART framed goals may be due to the fact that, in rehabilitation, the creation of SMART goals is often seen as difficult and time consuming (35), which may deter consistent use of the SMART framework for goal-setting by clinicians who face time constraints. Time has also been noted to be frequently missing from rehabilitation goals (16, 36). In line with these studies, many of the ambulatory goals within this study could not be qualified as SMART as they were missing a set time by which the goal is to be achieved. Clinicians may not document a set time frame, as the goal is assumed to be achieved by discharge.

Insufficient training and awareness among clinicians regarding the effective formulation and implementation of SMART goals may also impact the extent of its use. This is an especially prevalent issue when the definition of SMART goals varies, including variations for each of the acronyms like significant, meaningful, achievable, relevant and trackable that may be used to create the acronym SMART (15, 17). These variations in how clinicians interpret and apply the SMART framework could lead to inconsistencies in its use. This observation is consistent with prior research that underscores the need for standardized goal-setting practices to enhance adherence and outcomes in rehabilitation (21). It is also possible that other emerging goal setting measures such as ‘good goals’ and ‘goal-setting and action planning framework’ were used by the clinicians while setting goals instead of using the SMART framework (37). Additionally, due to the lack of consistent evidence supporting the use of SMART goals for improving patient outcomes (21), clinicians may find that the effort it takes to write SMART goals outweighs the possible benefits that it may provide, leading to its limited adoption.

This study was the first to examine the impact of SMART goals on ambulatory outcomes in outpatient stroke rehabilitation and the results demonstrate the importance of using a SMART goal format in rehabilitation. The finding that patients with SMART ambulatory goals demonstrated better ambulatory ability at discharge than those without SMART ambulatory goals, suggests that SMART goals may enhance the focus of rehabilitation efforts and facilitate progress monitoring.

The structured nature of SMART goals likely ensures that the goals set are aligned with the patient’s capabilities and needs, promoting better engagement and adherence to rehabilitation plans (10).

These findings align with existing evidence that well-defined, specific goals can improve treatment outcomes in neurological rehabilitation (12). Additionally, a previous study involving neurological patients showed that, while keeping their ultimate goal in mind, clients reported they were intrinsically motivated to work on specific goals like SMART goals to progress towards addressing their ultimate goal (38). Increased motivation to participate in rehabilitation is likely to lead to improved outcomes and goal attainment. *Canadian Stroke Best Practice Recommendations* indicate that stroke rehabilitation should involve interdisciplinary care (4), and SMART goals are thought to facilitate coordination between interprofessional team members to allow for the creation of individualized patient plans (4). This likely leads to improved care delivery and ultimately improved patient outcomes.

### Limitations

One of the limitations of the study is the potential variability in how physiotherapists interpret and use the SMART acronym. Although a consistent definition of the SMART acronym was applied within this study, physiotherapists may have used an alternate version of the SMART acronym that did not align with the definition used in this study to create a goal as there is no universally standardized version of each component of SMART. As a result, these alternate SMART goals may not have been captured within this study. Further evidence is needed to support and standardize the specific version of the SMART acronym used in this study. Additionally, as FAC measures ambulatory ability over short distances over various surfaces (25, 26), improvements in ambulatory endurance may not have been captured by this outcome measure. Furthermore, the study participants attended rehabilitation during the COVID-19 pandemic, which may have affected the availability and delivery of rehabilitation services, as well as patients’ access to healthcare facilities. These factors could have influenced the study outcomes and limit the generalizability of the findings to non-pandemic conditions.

### Implications for practice and future research

The study’s findings have important implications for clinical practice and future research. Enhancing clinician training and education in SMART goal-setting could improve the prevalence and consistent application of SMART goals in rehabilitation settings. Developing standardized protocols for goal-setting may help ensure that SMART goals are effectively implemented and monitored in order to improve goal attainment. As the results of this research indicate the effectiveness of SMART goals for improving ambulatory outcomes, future research is required to determine the effectiveness of SMART formatted goals on improving other functional outcomes (e.g., hand function). Further research is also warranted to investigate the mechanisms by which SMART goals influence patient adherence and progress.

## CONCLUSION

This study reveals a relatively low prevalence of SMART goals in outpatient stroke rehabilitation, with only 20.3% of patients having at least one SMART ambulatory goal documented. Although SMART goals had limited adoption, patients with SMART ambulatory goals experienced significantly better improvements in their FAC scores compared to those with non-SMART goals. The structured nature of SMART goals appears to enhance rehabilitation effectiveness by ensuring goals are specific, measurable, action-oriented, realistic and time bound. This may promote greater engagement and adherence to rehabilitation plans and allow for more effective progress monitoring. However, the generalizability of these findings may be constrained by the specific settings and population studied, which included three urban rehabilitation hospitals in Toronto. Variability in how physiotherapists interpret and apply the SMART framework, as well as potential differences in the definitions of SMART goals may also impact consistency in goal-setting practices. Future research should focus on developing an evidence-informed theoretical framework for setting SMART goals in rehabilitation settings to ensure consistent application and effectiveness (37).

Enhancing clinical training and developing standardized protocols for goal-setting could improve the prevalence and consistent application of SMART goals, potentially leading to better rehabilitation outcomes for stroke survivors.

## Data Availability

Individual-participant data are not available publicly due to privacy legislation.

## ACKNOWLEDGEMENTS

The authors would like to acknowledge David Jagroop, Kay-Ann Allen, Cynthia Danells, and Alex Kalli who collected the data. This research was funded by the Canadian Institutes of Health Research (PJT 173472).

## REFERENCES

1. Katan M, Luft AR. Global burden of stroke. Semin Neurol. 2018;38(2):208–11. doi: 10.1055/s-0038-1649503.

2. Holodinsky JK, Lindsay P, Yu AYX, Ganesh A, Joundi RA, Hill MD. Estimating the number of hospital or emergency department presentations for stroke in Canada. Can J Neurol Sci. 2023;50(6):820–5. doi: 10.1017/cjn.2022.338.

3. Yu AYX, Smith EE, Krahn M, Austin PC, Rashid M, Fang J, et al. Association of neighborhood-level material deprivation with health care costs and outcome after stroke. Neurology. 2021;97(15):e1503–11. doi: 10.1212/WNL.0000000000012676.

4. Teasell R, Salbach NM, Foley N, Mountain A, Cameron JI, de Jong A, et al. Canadian stroke best practice recommendations: rehabilitation, recovery, and community participation following stroke. Part one: rehabilitation and recovery following stroke; 6th edition update 2019. Int J Stroke. 2020;15(7):763–88. doi: 10.1177/1747493019897843.

5. Kapral MK, Fang J, Silver FL, Hall R, Stamplecoski M, O’Callaghan C, et al. Effect of a provincial system of stroke care delivery on stroke care and outcomes. Can Med J. 2013;185(10):E483–91. doi: 10.1503/cmaj.121418

6. Krueger H, Lindsay P, Cote R, Kapral MK, Kaczorowski J, Hill MD. Cost avoidance associated with optimal stroke care in Canada. Stroke. 2012;43(8):2198–026. doi: 10.1161/STROKEAHA.111.646091.

7. Heart and Stroke Foundation of Canada. Different strokes: recovery triumphs and challenges at any age. 2017.

8. Algurén B, Lundgren-Nilsson A, Sunnerhagen KS. Functioning of stroke survivors--a validiation of the ICF core set for stroke in Sweden. Disabil Rehabil. 2010;32(7):551–9. doi: 10.3109/09638280903186335.

9. Bohannon RW, Andrews AW, Smith MB. Rehabilitation goals of patients with hemiplegia. Int J Rehab Research. 1988;11(2):181–3.

10. Sugavanam T, Mead G, Bulley C, Donaghy M, van Wijck F. The effects and experiences of goal setting in stroke rehabilitation -a systematic review. Disabil Rehabil. 2013;35(3):177–90. doi: 10.3109/09638288.2012.690501.

11. Evensen J, Soberg HL, Sveen U, Hestad KA, Moore JL, Bronken BA. Individualized goals expressed by patients undergoing stroke rehabilitation: an observational study. J Rehabil Med. 2024;56:jrm15305. doi: 10.2340/jrm.v56.15305.

12. Rice DB, McIntyre A, Mirkowski M, Janzen J, Viana R, Britt E, et al. Patient-centred goal setting in a hospital-based outpatient stroke rehabilitation center. Phys Med Rehabil. 2017;9(9):856–65. doi: 10.1016/j.pmrj.2016.12.004.

13. Latham NK, Jette DU, Slavin M, Richards LG, Procino A, Smout RJ, et al. Physical therapy during stroke rehabilitation for people with different walking abilities. Arch Phys Med Rehabil. 2005;86(12 Suppl 2):S41–50. doi: 10.1016/j.apmr.2005.08.128.

14. Bailey RR. Goal setting and action planning for health behavior. Am J Lifestyle Med. 2017;13(6):615–8. doi: 10.1177/1559827617729634.

15. Wade DT. Goal setting in rehabilitation: an overview of what, why and how. Clin Rehabil. 2009;23:291–5. doi: 10.1177/0269215509103551.

16. Plant S, Tyson SF. A multicentre study of how goal-setting is practised during inpatient stroke rehabilitation. Clin Rehabil. 2018;32(3):263–72. doi: 10.1177/0269215517719485.

17. Lohmann S, Decker J, Müller M, Strobl R, Grill E. The ICF forms a useful framework for classifying individual patient goals in post-acute rehabilitation. J Rehabil Med. 2011;43(2):151–5. doi: 10.2340/16501977-0657.

18. Locke EA, Latham GP. New directions in goal-setting theory. Current Directions in Psychological Science. 2006;15(5):207–68. doi: 10.1111/j.1467-8721.2006.00449.x.

19. Janzen S, Mirkowski M, McIntyre A, Mehta S, Iruthayarajah J, Teasell R. Referral patterns of stroke rehabilitation inpatients to a model system of outpatient services in Ontario, Canada: a 7-year retrospective analysis. BMC Health Serv Res. 2019;19(1):399. doi: 10.1186/s12913-019-4236-5.

20. Rosewilliam S, Roskell CA, Pandyan AD. A systematic review and synthesis of the quantitative and qualitative evidence behind patient-centred goal setting in stroke rehabilitation. Clin Rehabil. 2011;25(6):501–14. doi: 10.1177/0269215510394467.

21. Levack WMM, Weatherall M, Hay-Smith EJC, Dean SG, McPherson K, Siegert RJ. Goal setting and strategies to enhance goal pursuit for adults with acquired disability participating in rehabilitation. Cochrane Database Syst Rev. 2015;2015(7):CD009727. doi: 10.1002/14651858.CD009727.pub2.

22. Preston E, Ada L, Stanton R, Mahendran N, Dean CM. Prediction of independent walking in people who are nonambulatory early after stroke: a systematic review. Stroke. 2021;52(10):3217–24. doi: 10.1161/STROKEAHA.120.032345.

23. Barzideh A, Devasahayam AJ, Tang A, Inness E, Marzolini S, Munce S, et al. Physiotherapists’ use of aerobic exercise during stroke rehabilitation: a qualitative study using chart-stimulated recall. medRxiv. 2023;doi:10.1101/2023.12.13.23299927. doi: 10.1101/2023.12.13.23299927.

24. Thompson S, Devasahayam AJ, Danells CJ, Jagroop D, Inness EL. Effect of cardiorespiratory exercise during rehabilitation on functional recovery early post-stroke: a cohort study. medRxiv. 2024;doi:10.1101/2024.08.09.24311772. doi: 10.1101/2024.08.09.24311772.

25. Holden MK, Gill KM, Magliozzi MR, Nathan J, Piehl-Baker L. Clinical gait assessment in the neurologically impaired: reliability and meaningfulness. Phys Ther. 1984;64(1):35–40. doi: 10.1093/ptj/64.1.35.

26. Mehrholz J, Wagner K, Rutte K, Meissner D, Pohl M. Predictive validity and responsiveness of the functional ambulation category in hemiparetic patients after stroke. Arch Phys Med Rehabil. 2007;88(10):1314–9. doi: 10.1016/j.apmr.2007.06.764.

27. College of Physiotherapists of Ontario. SMART learning goals [cited 2024 9 July]. Available from: https://www.collegept.org/docs/default-source/quality-assurance/qmf_smart_learninggoals_updated.pdf?sfvrsn=b889c9a1_0.

28. Gisev N, Bell JS, Chen TF. Interrater agreement and interater reliability: key concepts, approaches, and applications. Res Social Adm Pharm. 2013;9(3):330–8. doi: 10.1016/j.sapharm.2012.04.004.

29. Lohse K, Lang CE, Boyd LA. Is more better? Using metadata to explore dose-response relationships in stroke rehabilitation. Stroke. 2014;45:2053–8. doi: 10.1161/STROKEAHA.114.004695.

30. Dromerick AW, Geed S, Barth J, Brady K, Giannetti MK, Mitchell A, et al. Critical period after stroke study (CPASS): a phase II clinical trial testing an optimal time for motor recovery after stroke in humans. Proc Natl Acad Sci U S A. 2021;118(39). doi: 10.1073/pnas.2026676118.

31. Rohmann JL, Heurta-Gutierrez R, Audebert HJ, Kurth T, Piccininni M. Adjusted horizontal stacked bar graphs (“Grotta bars”) for consistent presentation of observational stroke study results. Eur Stroke J. 2023;8(1):370–9. doi: 10.1177/23969873221149464

32. Johns H. Convenient plotting for the modified Rankin scale and other ordinal outcome data. 1.1.0 ed2023.

33. Shieh G. Power analysis and sample size planning in ANCOVA designs. Psychometrika. 2020;85(1):101–20. doi: 10.1007/s11336-019-09692-3.

34. National Institute for Health and Care Excellence. Stroke rehabilitation in adults 2023 [cited 2024 16 July]. Available from: https://www.nice.org.uk/guidance/ng236.

35. Bovend’Eerdt TJH, Botell RE, Wade DT. Writing SMART rehabilitation goals and achieving goal attainment scaling: a practical guide. Clin Rehabil. 2009;23(4):352–61. doi: 10.1177/0269215508101741.

36. Tariah HA, Aljehani AS, Alenazi DY, Alturaif DA, Alsarhani MN. Occupational therapy goal achievement for people with stroke: a retrospective study. Occup Ther Int. 2020;2020:8587908. doi: 10.1155/2020/8587908.

37. Scobbie L, McLean D, Dixon D, Duncan E, Wyke S. Implementing a framework for goal setting in community based stroke rehabilitation: a process evaluation. BMC Health Services Research. 2013;13:190. doi: 10.1186/1472-6963-13-190.

38. Littooij E, Doodeman S, Holla J, Ouwerkerk M, Post L, Satink T, et al. Setting meaningful goals in rehabilitation: a qualitative study on the experiences of clients and clinicians in working with a practical tool. Clin Rehabil. 2022;36(3):415–28. doi: 10.1177/02692155211046463.

